# SARS-CoV-2 attack rate and population immunity in southern New England, March 2020 - May 2021

**DOI:** 10.1101/2021.12.06.21267375

**Authors:** Thu Nguyen-Anh Tran, Nathan B Wikle, Fuhan Yang, Haider Inam, Scott Leighow, Bethany Gentilesco, Philip Chan, Emmy Albert, Emily R Strong, Justin R Pritchard, William P Hanage, Ephraim M Hanks, Forrest W. Crawford, Maciej F Boni

## Abstract

Estimating an infectious disease attack rate requires inference on the number of reported symptomatic cases of a disease, the number of unreported symptomatic cases, and the number of asymptomatic infections. Population-level immunity can then be estimated as the attack rate plus the number of vaccine recipients who had not been previously infected; this requires an estimate of the fraction of vaccines that were distributed to seropositive individuals. To estimate attack rates and population immunity in southern New England, we fit a validated dynamic epidemiological model to case, clinical, and death data streams reported by Rhode Island, Massachusetts, and Connecticut for the first 15 months of the COVID-19 pandemic, from March 1 2020 to May 31 2021. This period includes the initial spring 2020 wave, the major winter wave of 2020-2021, and the lagging wave of lineage B.1.1.7(Alpha) infections during March-April 2021. In autumn 2020, SARS-CoV-2 population immunity (equal to the attack rate at that point) in southern New England was still below 15%, setting the stage for a large winter wave. After the roll-out of vaccines in early 2021, population immunity in many states was expected to approach 70% by spring 2021, with more than half of this immune population coming from vaccinations. Our population immunity estimates for May 31 2021 are 73.4% (95% CrI: 72.9% - 74.1%) for Rhode Island, 64.1% (95% CrI: 64.0% - 64.4%) for Connecticut, and 66.3% (95% CrI: 65.9% - 66.9%) for Massachusetts, indicating that >33% of southern Englanders were still susceptible to infection when the Delta variant began spreading in July 2021. Despite high vaccine coverage in these states, population immunity in summer 2021 was lower than planned due to 34% (Rhode Island), 25% (Connecticut), and 28% (Massachusetts) of vaccine distribution going to seropositive individuals. Future emergency-setting vaccination planning will likely have to consider over-vaccination as a strategy to ensure that high levels of population immunity are reached during the course of an ongoing epidemic.

## Introduction

Public health response and epidemic management of the COVID-19 pandemic met significant challenges at every stage of the pandemic in 2020 and 2021. Clinical experience and trial data accrued during the first and most deadly [1,2] wave of March-April 2020 leading to improvements in care for hospitalized patients [3–6]. Understanding of mobility, lockdown, and contact tracing policies improved in summer 2020, allowing for preparation of school reopening plans in autumn 2020 [7–9]. However, in fall 2020, substantial variation in results reported from several large seroprevalence studies [10–12] meant that we knew little at the time about the true number of individuals that had been infected between March 2020 and Nov 2020, and how strongly population susceptibility would drive the winter epidemic wave of 2020-2021.

Real-time estimation of seroprevalence or attack rate is challenging. A cross-sectional serological approach requires pre-planned periodic serum collections [13–15] and a high-throughput validated assay, but results will still be reported with a one month lag due to the delay from infection to IgG positivity in a serological assay. Estimates of attack rate using daily reported case numbers require us to be able to estimate (*i*) the number of unreported or untested symptomatic cases and (*ii*) the number of asymptomatic infections. In this estimation procedure, either an assumed infection fatality rate (IFR) [16,17], hospitalization incidence [1,18], or death incidence [19] can be used to work backwards to infer the numbers of unreported cases or unreported infections. This means that age structure is necessary in these reporting streams, as the rate of asymptomatic SARS-CoV-2 infection, hospitalization probability, and death probability all vary substantially by age [20–22]. When hospitalization incidence is not available (e.g. due to under-reporting [1]), data streams for death, current hospitalization, current numbers of patients in intensive care and on ventilators can be used, with some degree of accuracy, to estimate the incidence of hospitalization. Knowing the duration of a typical medical-floor hospital stay improves the estimates.

Since the beginning of the COVID-19 epidemic in the US, the Centers for Disease Control and Prevention (CDC) have been collecting cross-sectional serum samples, both from blood donors and residual samples from routine laboratory testing [10,12]. These sample collections are a valuable epidemiological resource, but for the majority of states seroprevalence estimates using these samples do not seem to be translatable into attack rate estimates because the seroprevalence estimates move and up and down through time while the attack rate can only go up [23]. These non-monotonic measurements are common in serology; if an antibody assay threshold is set too high the assay shows ‘recent seroprevalence’ rather than ‘seroprevalence’, resulting in systematic under-estimation of the number of individuals that have been infected. A simple example can be seen for Massachusetts infection seroprevalence, measured as 10.2% in late April 2021 [24], at which point 9.1% of the state’s residents had reported as a confirmed positive COVID-19 case. This would mean that only about 11% of SARS-CoV-2 infections in Massachusetts were asymptomatic or unreported, which is inconsistent with our knowledge of SARS-CoV-2 clinical progression and fatality rates.

In this analysis, we present a model-based reconstruction of the SARS-CoV-2 attack rate and population-immunity curves for three New England states – Massachusetts (MA), Connecticut (CT), and Rhode Island (RI) – for the first 15 months of the pandemic. We include the 2021 vaccination campaign and estimate the fractions of the vaccine supply that were distributed to seropositive and seronegative individuals. This allows us to estimate the fraction of each state’s population that was immune by May 31 2021, before the SARS-CoV-2 Delta variant arrived in the northeast in summer 2021 causing an upturn in cases that began in late July 2021. Using our attack-rate estimates, we estimate the infection fatality rates in all three states and show, as in previous analyses [1,2], that the March/April 2020 wave was deadlier on a per-infection basis than subsequent epidemic waves. In addition, we show that a higher than expected percentage of the vaccine supply was apportioned to individuals that had already been infected with SARS-CoV-2, lessening the positive impact of the winter/spring 2021 vaccination campaign.

## Methods

A validated Bayesian inferential framework based on a dynamical epidemic model (compartmental diagram in Supplementary Figure 1) was used to fit case, hospitalization, and death data from Massachusetts, Connecticut, and Rhode Island [1,25]. Eleven daily data streams were collected from each state: (1) cumulative confirmed cases, (2) cumulative confirmed cases by age, (3) cumulative hospitalized cases, (4) cumulative hospitalized cases by age, (5) number of patients currently hospitalized, (6) number of patients currently in ICU, (7) number of patients currently on mechanical ventilation, (8) cumulative deaths, (9) cumulative deaths by age, (10) cumulative hospital deaths, (11) cumulative hospital discharges. Note that the age-stratified data do not always sum to the all-ages data streams. Daily time points from March 1 2020 to June 6 2021 were included in this analysis. Cumulative hospitalization data were included for MA even though these data were not available for our previous two analyses [1,25]. Details on CT data sources are in the Supplementary Methods; RI and MA data sources are described in Wikle et al [1].

Vaccination campaigns were added into the dynamic model. Weekly age-structured SARS-CoV-2 vaccination numbers were obtained separately for the three states [26–29], and vaccinated individuals were moved from the susceptible compartment in the dynamical model to the recovered compartment whenever a seronegative individual was vaccinated. The model allows for vaccination of seropositive individuals. In the model, the fraction

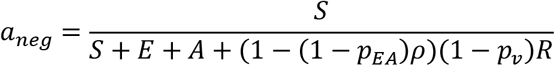

represents the total fraction on non-symptomatic non-hospitalized individuals that are currently antibody negative and virus negative. Above, the denominator is the candidate pool of individuals for whom COVID vaccine would be recommended. The uppercase letters represent the compartments in the model: the current number of susceptible individuals (*S*), the number exposed (*E*), the number asymptomatically infected (*A*), and the number recovered from a non-hospitalized infection or who are post-vaccination (*R*). The fraction *p*_*EA*_ represents the fraction of infected individuals that progress to asymptomatic infection (different for every age group), and the reporting rate *ρ* represents the fraction of symptomatic individuals that were tested, confirmed positive, and are aware that they have already had COVID-19. The fraction *p*_*v*_ is the total fraction of the population (by age group) that has been vaccinated thus far. Thus, the denominator’s modified *R*-term seeks to approximate the non-vaccinated fraction of the recovered group who are unaware that they have recovered from a SARS-CoV-2 infection, and thus may have sought vaccination between January and May 2021. The model reports the total number of vaccines given to individuals in the susceptible class *S*, and the total number of vaccines given to individuals in all classes.

The model accommodates temporally varying patterns of clinical care and changing age-contact rates. To model either a change in clinical management or an increase/decrease in the vulnerability of the current patient pool, we allow the transition probability from medical-floor stay to intensive care unit (ICU) to vary throughout the epidemic. We used the age-specific probabilities of progression to ICU from Lewnard et al [20] and scaled them (simultaneously, to keep the relative probabilities the same across ages) to allow for three separate severity or vulnerability periods during the 15-month epidemic. Three periods of age-contact patterns were allowed for CT and RI, and four were used for MA (based on lower DIC and visual fit). Wide priors were set for the timings of these transitions; see Supplementary Materials.

The reporting rate – the fraction of symptomatic cases that are reported to state-level health systems – was modeled for each state with an I-spline (a non-decreasing function). Symptomatic infections counted as unreported are those where the individual (*i*) chose to stay home without reporting to a health provider or testing center, or (*ii*) did not receive a SARS-CoV-2 PCR test for any other reason. Attack-rate estimates are obtained as the sum of reported symptomatic cases, unreported symptomatic cases, and asymptomatic infections (using external estimates from Davies et al [22]).

We report two types of infection fatality rates (IFR). The population-weighted infection fatality rate (pIFR) is the probability of death, if infected, for a person sampled at random from a population with a particular age structure. The epidemic infection fatality rate (eIFR) is infections weighted; it is the probability of death for a randomly sampled infected individual during a particular epidemic phase. All estimates are presented as medians and credible intervals from 1000 posterior samples.

## Results

State-level inference shows model fits that accurately describe the dynamics of ICU occupancy, ventilator occupancy, and daily deaths counts in all three states during the study period. The Rhode Island inferred epidemic curve in particular shows close fits to all eight non-age structured data streams (Figure 1), likely due to the completeness of hospital reporting available in a small state. Both the data and the model – across all data streams, for all three states – clearly reconstruct the early epidemic wave of March-April 2020, the summer lull of 2020, the major winter wave of 2020-2021, and the lagging wave of the Alpha (B.1.1.7) variant in March-April 2021. Case and hospitalization data were fit well in all states with the exception of the case incidence data and current hospitalization data in Massachusetts which the model under-fit for the March-April 2020 epidemic wave (Supplementary Figure 2). In addition, high variance in new case incidence in Connecticut for the March-April 2020 wave and the major winter wave of 2020-2021 suggest that the model may not be capturing complete heterogeneity in transmission dynamics and case reporting (Supplementary Figure 3).

**Figure 1.**
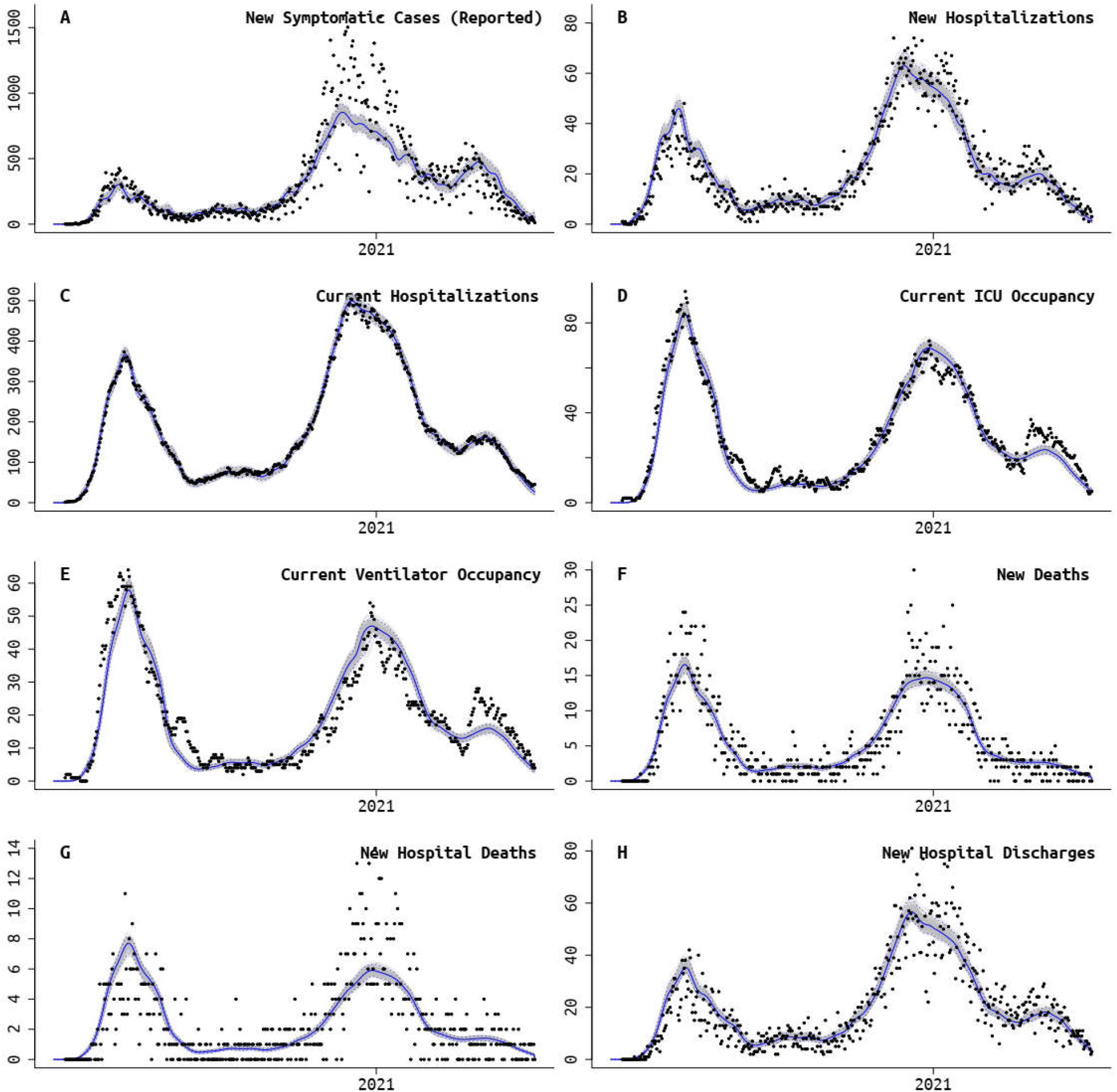
Rhode Island fit of model to data. Panels **A, B**, and **F** also have age-structured data streams, making a total of 11 data streams that were fit. Black dots are absolute daily counts. Blue line is model median from the posterior, and gray bands show 95% credible region.

Using our model’s inferred reporting rate and an external estimate [22] of the asymptomatic fraction of each age group’s infections, we inferred that as of May 31 2021 the population-level attack rates were 41.5% in RI (95% CrI: 40.4% - 42.7%), 25.8% in CT (95% CrI: 25.5% - 26.3%), and 28.0% in MA (95% CrI: 27.1% - 29.0%). Since summer 2020, attack rate estimation has been robust to the differing amounts of data included in the analyses (Figure 2). Attack-rate estimates in Connecticut are consistent with those reported in Morozova et al [30], and attack-rate comparisons in Rhode Island and Massachusetts are consistent with other model-based estimates [16,17] as described in Wikle et al [1]. Vaccination began in all three states in December 2020, with an initial rollout that was slower than expected. By January 31 2021, approximately 2.1% to 2.3% of each state’s population was vaccinated, with this vaccinated fraction reaching 8.0% to 9.6% by February 28 2021. Using (*i*) the modeled number of infections, (*ii*) daily data on vaccinations that were integrated into the model, and (*iii*) the modeled number of seropositives that would have received vaccination (to remove double-counting of individuals who were naturally immune and then vaccinated), we infer population-immunity estimates of 73.4% in Rhode Island (95% CrI: 72.9% - 74.1%), 64.1% in Connecticut (95% CrI: 64.0% - 64.4%), and 66.3% in Massachusetts (95% CrI: 65.9% - 66.9%) for May 31 2021; see Figure 3. This implies that >33% of southern New England was immunologically naive when the Delta variant reignited a wave of infections in late July 2021. From the model, we infer that the percentage of vaccines administered to seropositive individuals was 34.1% in RI (95% CrI: 32.9% - 35.2%), 24.6% in CT (95% CrI: 24.3% - 25.1%), and 27.6% in MA (95% CrI: 26.8% - 28.6%). See Table 1 for a breakdown of infection and vaccination status in all three states; these estimates are consistent with those of Moghadas et al [31] who used a direct IFR-based deaths-to-infections translation to estimate that approximately half of previously infected individuals received vaccination.

**Figure 2.**
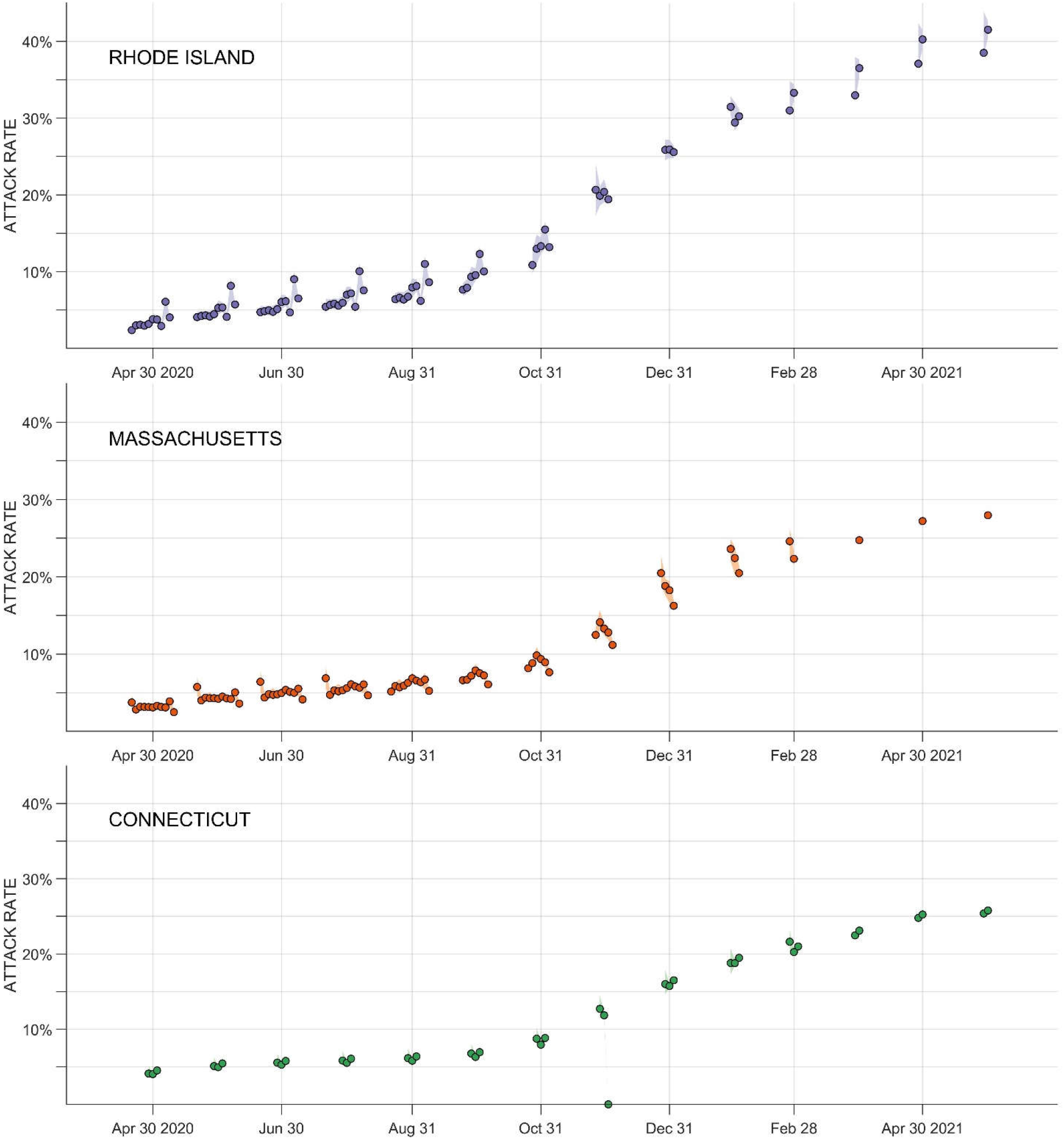
Each dot shows one attack-rate estimated with data available only through a particular date. For example, for April 30 2020, 10 estimates are available for Rhode Island, 11 estimates are available for Massachusetts, and three estimates are available for Connecticut; all of these estimates were obtained at different times with different amounts of data available. The dots are ordered from left to right chronologically, with the right-most estimates using the most data (and being done the latest). Shaded areas – sometimes too small to be seen – show 95% credible intervals for each estimate.

**Figure 3.**
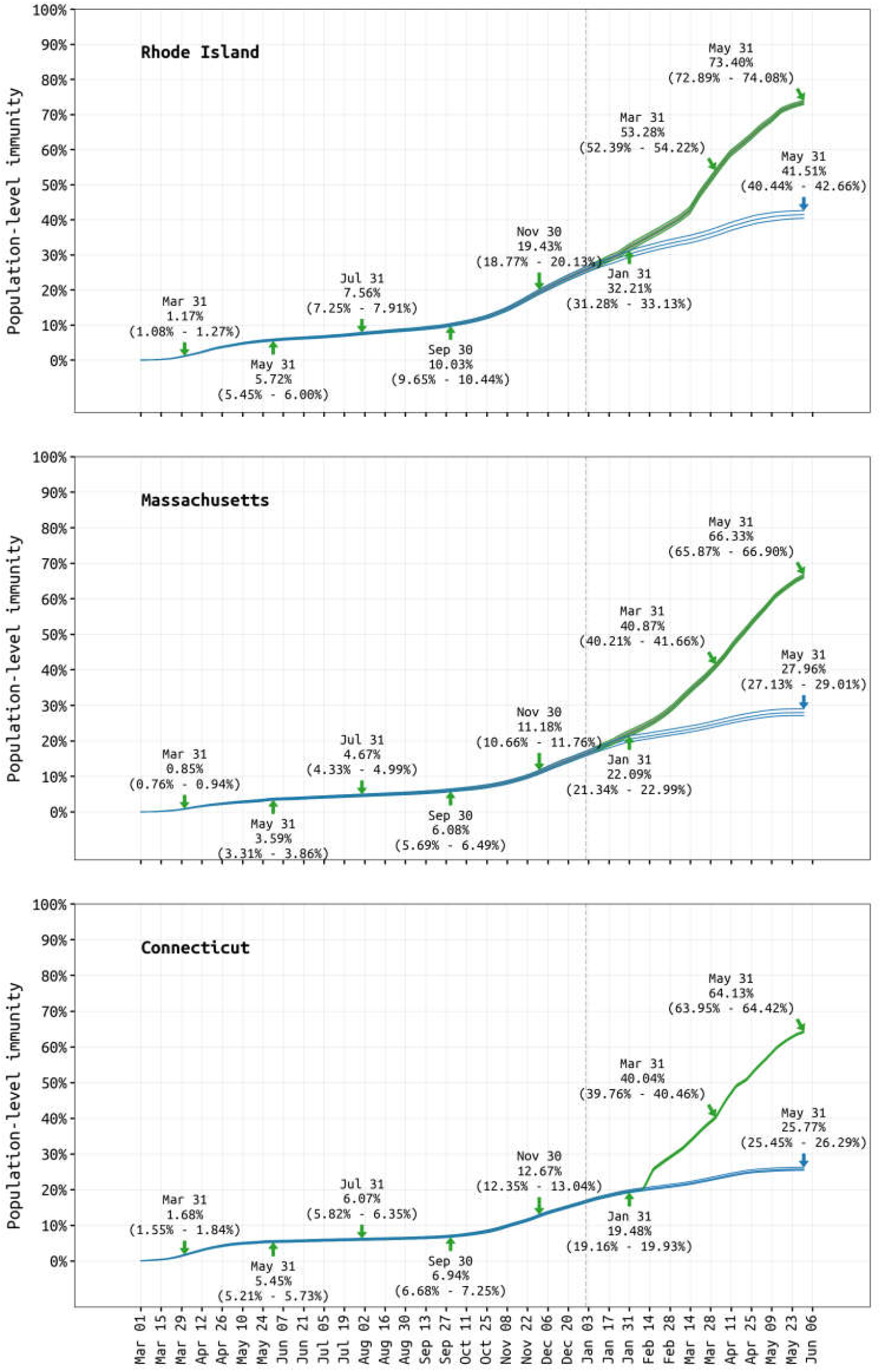
Blue lines show total percentage of each state’s population that has been infected. Green lines show percentage of the population that has either been infected or vaccinated (counting only once individuals who have been both infected and vaccinated). Three lines shown are median estimates and boundaries of 95% credible interval. Exact estimates shown every two months.

**Table 1.**
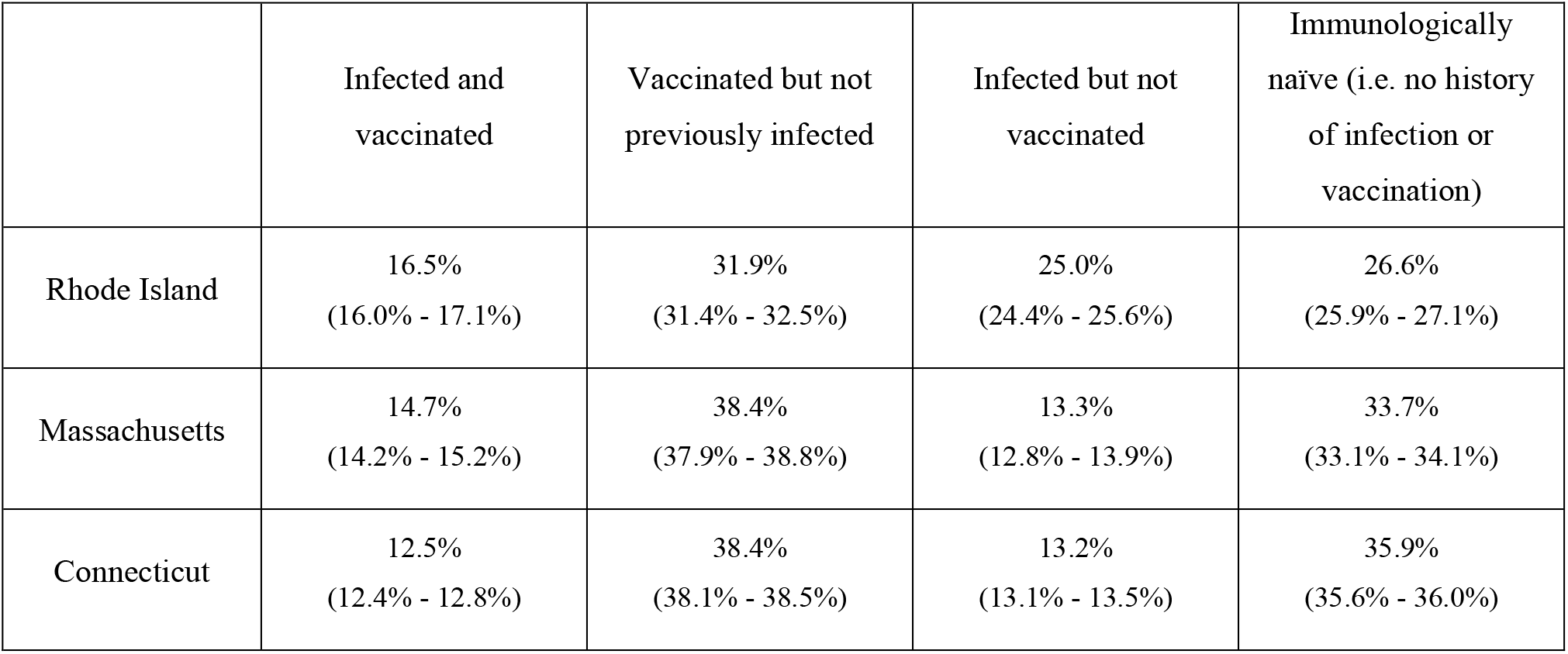
As of May 31 2021, inferred percentages (95% credible intervals) of each state’s population with particular vaccination status and prior infection status.

The mortality impact of the SARS-CoV-2 epidemic in southern New England was severe. In Connecticut, 0.229% of residents died over the first 15 months of the epidemic; 0.248% of RI residents and 0.249% of MA residents died over this same time period, indicating that the epidemic was around 8% or 9% more deadly in RI and MA than in CT. Using the inferred attack rates over the first 15 months of the epidemic, we estimate the 15-month eIFR in the three states as 0.62% in RI (95% CrI:0.60% - 0.64%), 0.89% in CT (95% CrI: 0.87% - 0.90%), and 0.89% in MA (95% CrI:0.86% - 0.92%). Rhode Island had an estimated 55% to 60% more infections per population than CT or MA, and this cannot be explained by any age-specific differences in transmission, indicating that Rhode Island had a larger and broader epidemic across all age groups. The lower eIFR in Rhode Island suggests that the larger epidemic extended to less vulnerable groups, lowering the average fatality rate for the epidemic as a whole; see *Discussion*.

As in our previous analyses on population-level signals showing evidence of different patterns of clinical progression during different epidemic phases [1,25], we include changepoints in the ICU admission fraction in our model to allow for changes in clinical management for hospitalized patients; a lower ICU admission fraction suggests that hospitalized patients have improved chances of recovery and a lower chance of death. The first changepoint was inferred as Jun 2 for RI, May 26 for MA, and Jun 5 for CT (medians from posteriors), and the second changepoints were inferred as Dec 10 for RI, Sep 12 for MA, and Nov 6 for CT (see Supplementary Materials for all posteriors). In Rhode Island and Massachusetts, the ICU admission fraction dropped substantially from the March-April 2020 epidemic wave to the summer/fall transmission period in 2020; Connecticut estimates may be less reliable as ICU data only began to be reported in July 2020. In early summer 2020, the age-adjusted probability of ICU admission dropped from 0.21 to 0.09 in Rhode Island, 0.45 to 0.17 in Massachusetts, and 0.20 to 0.17 in Connecticut; see Figure 4B.

**Figure 4.**
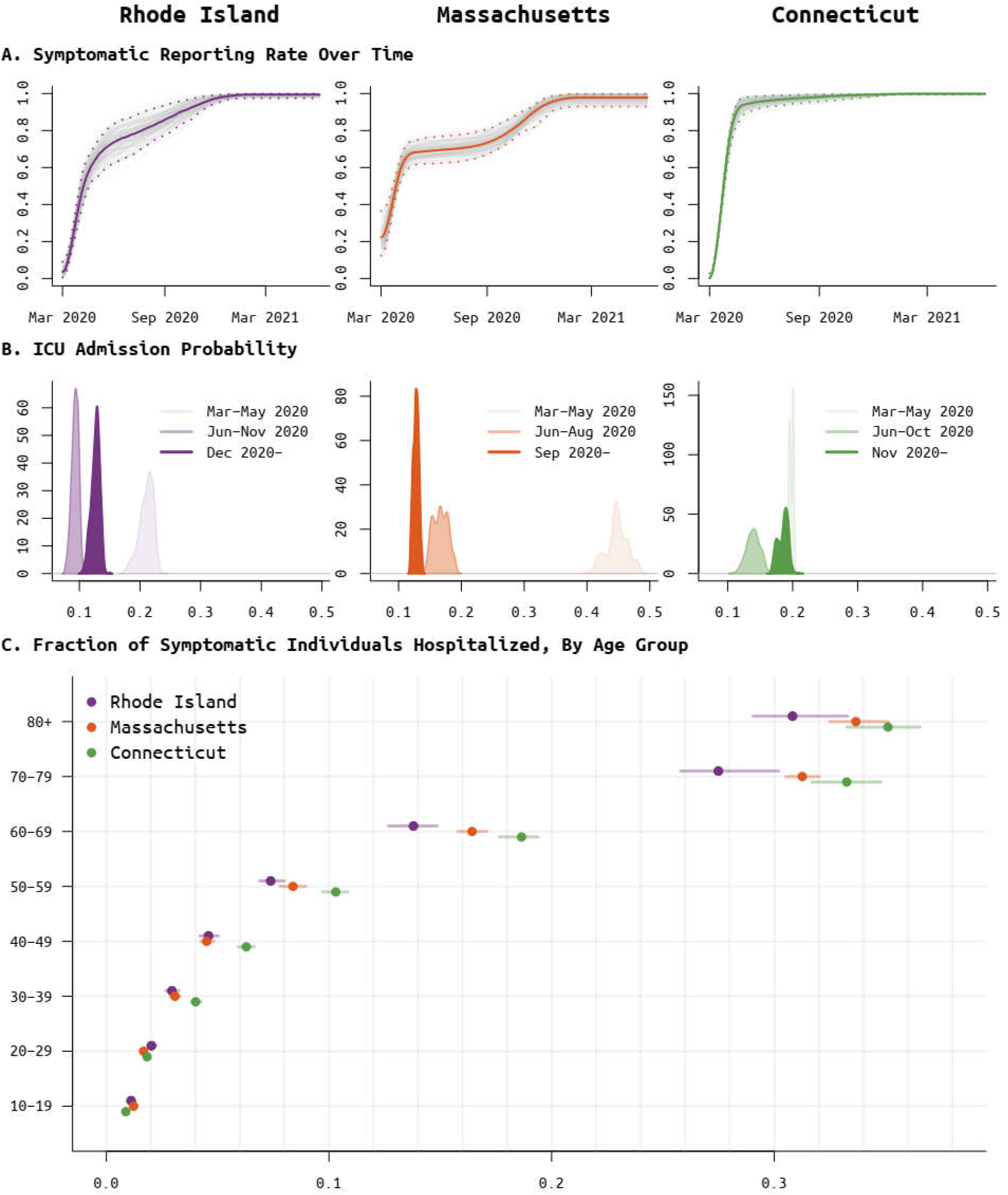
Posterior distributions for the (**A**) per-symptomatic-case reporting rate, (**B**) ICU admission probability, per hospitalized case, for different phases of the epidemic, and (**C**) the age-specific probability of hospitalization for symptomatic cases.

The estimated population-weighted pIFR estimate for the first phase of the epidemic was 1.64% for Rhode Island (95% CrI:1.52% - 1.72%), 1.55% for Connecticut (95% CrI:1.45% - 1.62%), and 2.40% for Massachusetts (95% CrI:2.22% - 2.51%), about two to three times higher than during later phases of the pandemic. This is consistent with previous analyses showing higher fatality rates in the earliest epidemic period which was characterized by substantial transmission in congregate care settings and no available clinical interventions (e.g. corticosteroids [5,6], prone positioning [3], antivirals [4]) that would improve survival odds for a hospitalized patients.

## Discussion

The unpredictability and periodic evolutionary jumps of the SARS-CoV-2 epidemic have made parts of public health planning impossible. Our inability to see several months ahead was partially caused by our inability to assess the amount of infection and immunity in the population at present. This was the reason that the epidemiology community did not foresee the beginning of the Delta wave in July 2021 [32], because we did not have accurate state-level estimates of population susceptibility. This had important implications as the lack of epidemiological preparations resulted in 140,000 individuals dying in the US during the Delta-epidemic period that lasted from July to October. In addition to lacking a system for real-time attack-rate estimation, we did not have an approach for co-analyzing the waning epidemic dynamics of January to May 2021 with the vaccine roll-out that was occurring simultaneously. As a result, 27% of vaccines in southern New England were administered to antibody-positive individuals, leading to overconfidence in the vaccination rollout’s effect on generating population immunity and preventing future waves of infection. As a matter of policy, for the next emergency-initiated vaccination campaign it will be necessary to consider over-vaccination as an option to ensure that we are not attempting a “soft landing” [33,34] with just enough vaccine distribution to reach an uncertain threshold of population immunity.

Despite the large volume of data that is now available for county-level and state-level case counts and hospital counts for the SARS-COV-2 epidemic in the United States [35], estimates of attack rate are not readily computable at these geographic levels. This shortcoming became obvious in July 2021, as accurate estimates of the attack rate would have been invaluable in assessing whether certain state populations had enough remaining susceptibility to support an incoming wave of the Delta variant which was imported to the US sometime in late spring 2021 [36]. Here, we demonstrate that live attack-rate estimation is possible from a combination of case and hospitalization data as these can be used to estimate the underreporting factor for symptomatic infections [1,18]. We combine eleven data streams for this analysis. We validate our model-fitting approach by showing that it has excellent visual fits to the most reliable data types (ICU counts, ventilator counts, and death counts), and that attack-rate estimates are robust as more data are added to the case time series every month (Figure 2).

Fitting age-structured transmission models to multiple data streams of cases, hospitalizations, ICU occupancy, ventilator occupancy, and deaths does present challenges that limit some of our interpretations. The exact rates and probabilities in a patient’s clinical progression path are not fully known (Supplementary Figure 1). Some patients will be more vulnerable to severe clinical outcomes, while other groups of patients may report late in their course of disease resulting in an apparent accelerated progression from presentation to hospitalization or death. When patient characteristics like these are not known, a transmission model estimates average rates and average probabilities for different age groups, hiding the variability that exists within each age group or state. Thus, if some state’s epidemic was biased towards healthier individuals, individuals with better access to care, or more vulnerable groups of individuals, our inferential model would not be able to detect these signals.

In our analysis, the three state epidemic profiles differ. Rhode Island’s epidemic is larger, but less severe on a per-infection basis. The epidemic in Massachusetts has different patterns of severity and hospital admission than the other two states. One potential explanation for the Rhode Island epidemic profile is a positive correlation between susceptibility and vulnerability. During an epidemic in a heterogeneous population, the most susceptible individuals are infected first [37,38]. This would mean that in the larger RI epidemic, the average susceptibility and the average vulnerability would be lower than in MA or CT, resulting in fewer hospitalizations per infection and a lower IFR in Rhode Island. For Massachusetts, it is likely that cumulative hospitalization counts are under-reported as these are self-reported by hospitals (personal communication, MA Dept of Public Health). If hospitalization incidence is undercounted in Massachusetts by 30%, then MA and CT would have nearly identical epidemic profiles with 9.4% of the population symptomatically infected, 0.87% of the population hospitalized, and a hospital fatality rate in Massachusetts that is about 10% higher than in Connecticut. This could be one of the reasons for the discrepancy between the hospital incidence (Supplementary Figure 2B) and current hospitalization data stream (Supplementary Figure 2C) in Massachusetts. A second possibility is an inability of the model to capture the long hospitalization times seen in MA during March-April 2020.

The most important information to integrate into the next phase of attack-rate estimation and population-immunity estimation in the US is the rate of antibody waning following SARS-CoV-2 infection and SARS-CoV-2 vaccination. Average waning rates are now known for the short-term post-infection [39– 42] and post-vaccination [43–46][47], and this may be enough to estimate a recent attack rate [48] which can either be reported as such or chained together with other recent attack-rate estimates to provide an annual attack-rate estimate. Although the initial live integration of these data streams will no doubt be challenging, the benefit will be a situationally-aware susceptibility estimate that will allow us to evaluate the invasion ability of a new high-transmissibility variants (such as Alpha or Delta) or the inevitable immune-escape variants – like the Omicron lineage [49,50] – that will likely continue appearing in 2022 and 2023. For Omicron specifically, the re-infection hazards presented by Pulliam et al [51] indicate that the “infected but not vaccinated” portion of the population presented in Table 1 can now be viewed as approximately twice as likely to be infected during the coming Omicron wave than they were in the previous Delta and Alpha waves. The cost of not providing these live attack-rate and susceptibility estimates is a repeat of summer 2021 when epidemiologists were caught unaware of the immediate risk posed by the introduction of the Delta variant in a still highly susceptible United States population.

The real-time exercise organized for the purpose of providing these monthly attack-rate estimates (http://mol.ax/covid) shows the value of understanding an epidemic’s susceptibility curve while it is changing and the importance of continuous IFR estimation as the epidemic passes through different vulnerability strata during different time periods. The work presented here shows that the population-weighted IFR in the earliest phases of the epidemic was much higher – between 1.4% and 2.8% – than the IFR estimates known at the time, that highly vaccinated populations were susceptible to a surge of Delta infections in July 2021, and that we over-estimated the spring 2021 vaccination campaign’s effect on population immunity. Both live attack-rate estimation and continuous IFR estimation can be sharpened by the addition of a data stream that connects case numbers to a whole-population measure. The most direct approach to this is to perform weekly PCR-testing on random samples of the population (or a cohort) to obtain basic live prevalence curves during an epidemic [52]. Knowing that tools like this are affordable and easily integrated into sample processing pipelines and data analysis pipelines should motivate us to include live attack-rate and susceptibility estimation into preparation plans for our next major uncontrolled epidemic.

## Data Availability

All data and code are available at https://github.com/bonilab/covid19-attackrate-RI-MA-CT

https://www.medrxiv.org/content/medrxiv/early/2021/08/18/2020.11.17.20232918/DC1/embed/media-1.pdf?download=true

## Acknowledgments

Thanks to Larry Madoff and Catherine Brown at the Massachusetts Department of Public Health for help in interpretation of the COVID-19 epidemic in Massachusetts.

## Author Contributions

M.F.B. and T.N-A.T. conceived the study. T.N-A.T. and N.B.W. performed the analysis. F.Y. and F.W.C. assembled the Connecticut COVID-19 data and validated the CT model fits. H.I. assembled and cleaned an updated version Massachusetts case and hospitalization data. All authors commented on the final results and edited the manuscript. M.F.B. and T.N-A.T. drafted the manuscript.

## Data and Code Availability

All data and code publicly available at https://github.com/bonilab/covid19-attackrate-RI-MA-CT.

