## Supplementary Materials for "SARS-CoV-2 attack rate and population immunity in southern New England, March 2020 - May 2021"

**Supplementary Methods***Collection of Massachusetts and Rhode Island data streams*

Data streams for MA and RI were collected as outlined in Wikle et al (*Sci Adv*, in press, 2021). In addition, age-structured cumulative hospitalization incidence was added for MA as this data stream was not available for the previous analysis. Details can be seen by downloading the pre-print PDF supplement at this link: https://www.medrxiv.org/content/medrxiv/early/2021/08/18/2020.11.17.20232918/DC1/embed/media-1.pdf?download=true.

*Aggregation and cleaning of Connecticut data streams*

Eleven data streams were sought in the Connecticut Department of Public Health data: (1) cumulative confirmed cases, (2) cumulative confirmed cases by age, (3) cumulative hospitalized cases, (4) cumulative hospitalized cases by age, (5) number of patients currently hospitalized, (6) number of patients currently in ICU, (7) number of patients currently on mechanical ventilation, (8) cumulative deaths, (9) cumulative deaths by age, (10) cumulative hospital deaths, (11) cumulative hospital discharges. Data streams (1) and (8) were collected from daily test results of CT DPH (https://data.ct.gov/Health-and-Human-Services/COVID-19-daily-and-cumulative-cases-deaths-and-tes/5dch-cm68). Cases were defined as laboratory-confirmed positive COVID-19 tests. Currently hospitalized patient counts were collected from the daily reports of CT DPH (https://data.ct.gov/Health-and-Human-Services/COVID-19-Tests-Cases-Hospitalizations-and-Deaths-S/rf3k-f8fg). Age-stratified cumulative cases and deaths came from the CT DPH (https://data.ct.gov/Health-and-Human-Services/COVID-19-Cases-and-Deaths-by-Age-Group/ypz6-8qyf). The remaining data streams were collected from other sources. Current ICU cases were collected from the hospital utilization report of Health and Human Services (https://healthdata.gov/Hospital/COVID-19-Reported-Patient-Impact-and-Hospital-Capa/g62h-syeh) and are defined as the number of adult patients currently hospitalized in an ICU bed. Cumulative hospital discharges were obtained from the COVID Tracking Project (https://covidtracking.com/data/state/connecticut). Total and age-stratified cumulative hospitalized cases were calculated from the cumulative hospitalization rates from COVID-NET

(https://gis.cdc.gov/grasp/COVIDNet/COVID19_3.html) and the population estimates from 2019 US census (https://www.census.gov/quickfacts/fact/table/US/PST045219).

##### *Vaccination Data in Connecticut*

Total (https://data.ct.gov/Health-and-Human-Services/COVID-19-Vaccination-Status-by-Residence-in-a-SVI-/tttv-egb7) and age-stratified fully vaccinated population numbers (https://data.ct.gov/Health-and-Human-Services/COVID-19-Vaccinations-by-Age-Group/vjim-iz5e) were collected from the CT DPH website. A person is considered fully vaccinated if they received two doses of the Pfizer or Moderna vaccines or one dose of the Johnson & Johnson vaccine.

### Supplementary Figures

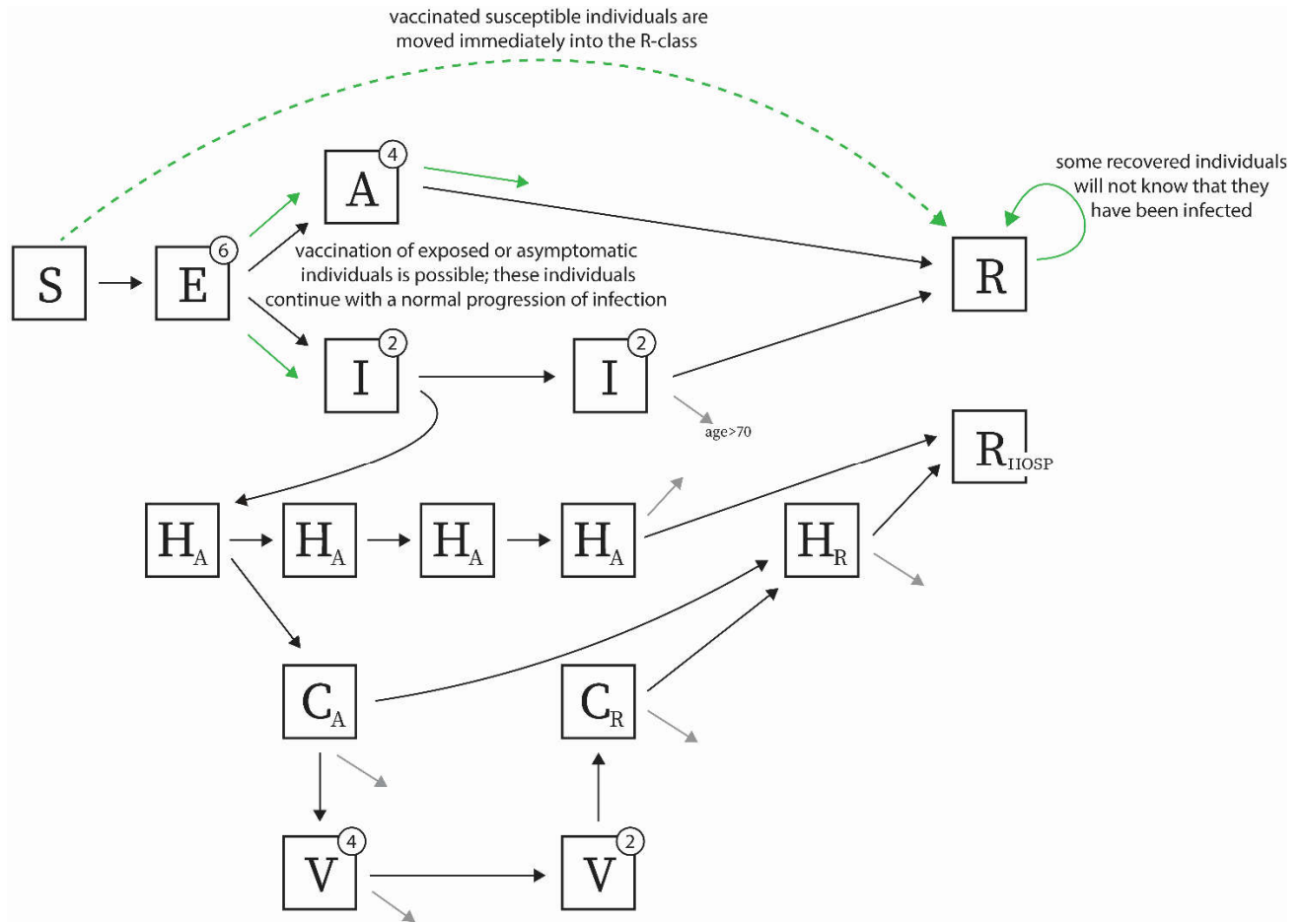

**Supplementary Figure 1.** Compartmental diagram for ordinary differential equations (ODE) epidemic model used for inference. Each compartment is broken down into nine 10-year age bands. Individuals can be susceptible (S), exposed (E), asymptomatic (A), infected and symptomatic but not hospitalized (I), hospitalized in the acute stage of infection (H<sub>A</sub>), in critical care during acute phase (C<sub>A</sub>), on a ventilator (V), in critical care after being removed from mechanical ventilation (C<sub>R</sub>), hospitalized and convalescing after being discharged from the ICU (H<sub>R</sub>), recovered from non-hospitalized infection (R), recovered from an infection that required hospitalization (R<sub>HOSP</sub>).

Numbers in upper-right corner of each compartment show number of transitory states used to model that patient group. For example, there are six exposed classes, E<sub>1</sub> to E<sub>6</sub>, each lasting approximately one day.

Gray arrows indicate death.

Green arrows show vaccination, but only the dashed green arrow moves individuals from one state to another.

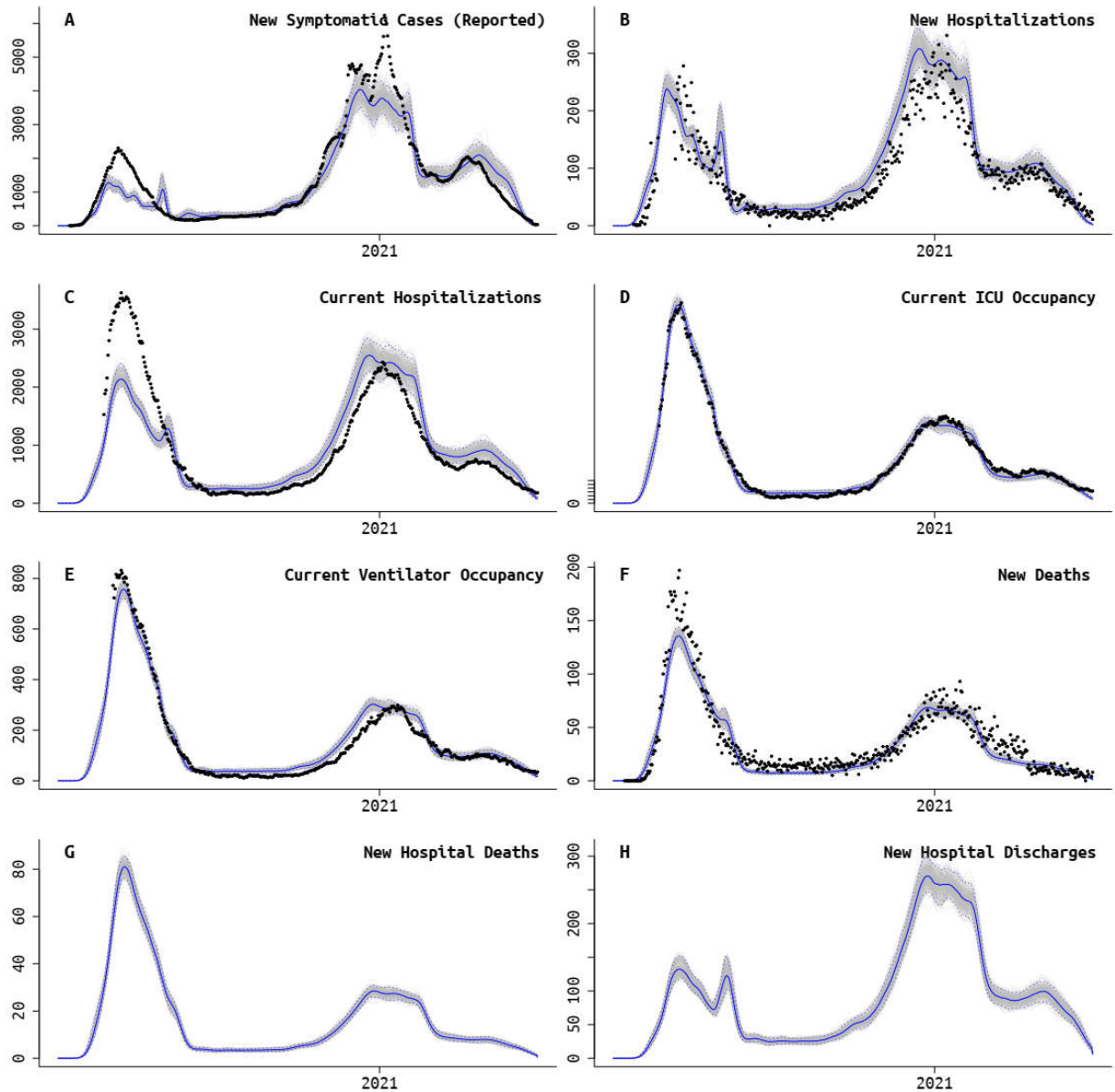

**Supplementary Figure 2.** Massachusetts fit of model to data. Panels **A**, **B**, and **F** also have age-structured data streams. Hospital discharge data and death data separated by in/out of hospital were not available in Massachusetts. Black dots are absolute daily counts. Blue line is model median from the posterior, and gray bands show 95% credible region.

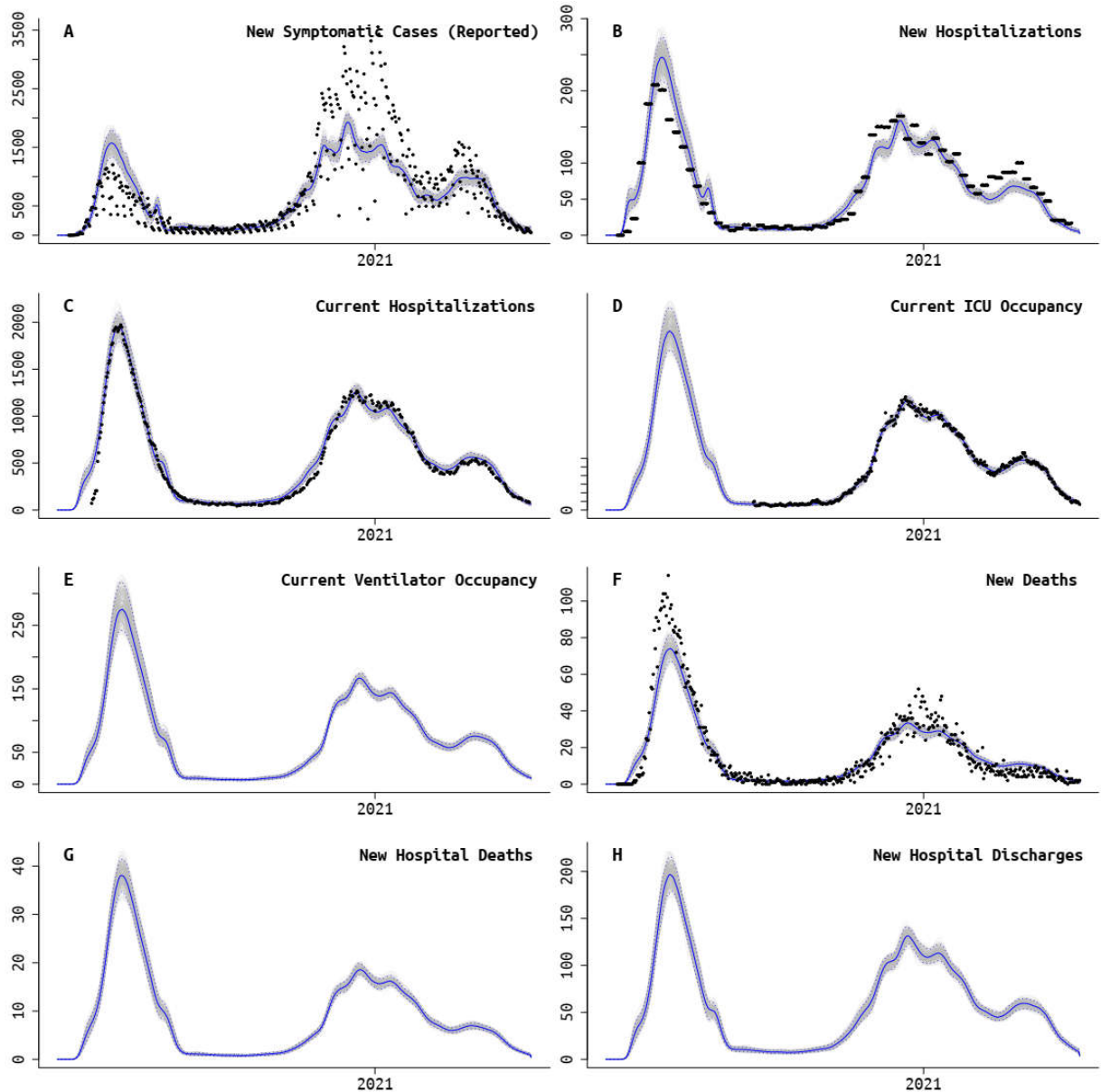

**Supplementary Figure 3.** Connecticut fit of model to data. Panels A, B, and F also have age-structured data streams. Hospital discharge data, death data separated by in/out of hospital, and ventilated patient counts were not available in Connecticut. Black dots are absolute daily counts. Blue line is model median from the posterior, and gray bands show 95% credible region.

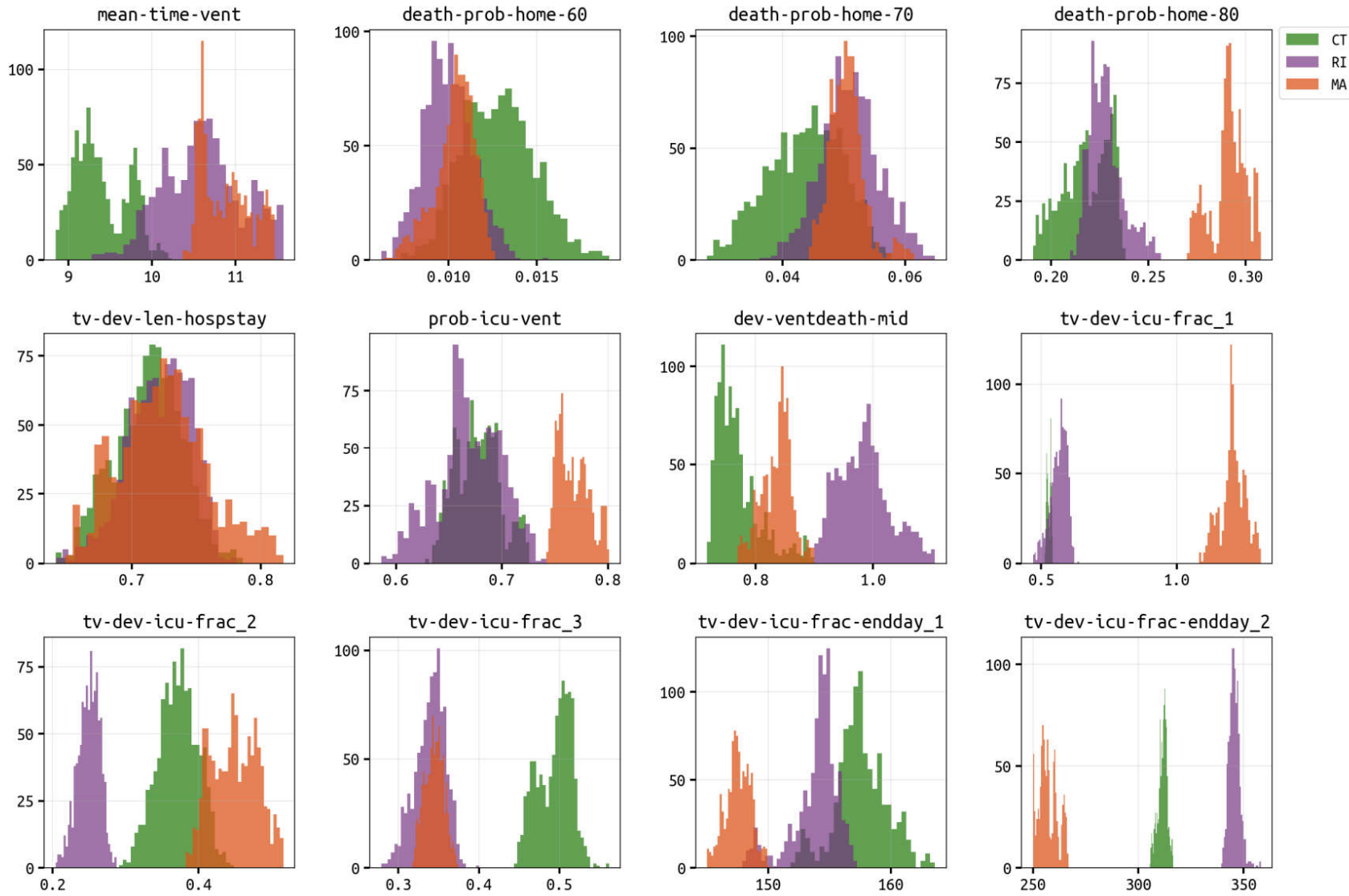

**Supplementary Figure 4.** Panels show posterior distributions for mean number of days a patient spends on a ventilator (**mean-time-vent**); age-specific probability of death in ten-year age bands for symptomatic patients outside hospital settings (e.g. **death-prob-home-60** for the 60-69 age group); the average length of non-ICU hospital stay (**tv-dev-len-hospstay**, multiply by 10.8 days); probability of progression from non-ventilated to ventilated status in the ICU (**prob-icu-vent**); deviation from expected mortality rate for 40-70 year-olds on ventilators (**dev-ventdeath-mid**); for three periods of the epidemic, the relative probability of ICU admission for hospitalized patients (**tv-dev-icu-frac\_1**, **tv-dev-icu-frac\_2**, **tv-dev-icu-frac\_3**); the end-days of the first two periods with aforementioned ICU admission probabilities (**tv-dev-icu-endday\_1**, **tv-dev-icu-endday\_2**, day 150 is May 30 2020, day 300 is Oct 26 2020).

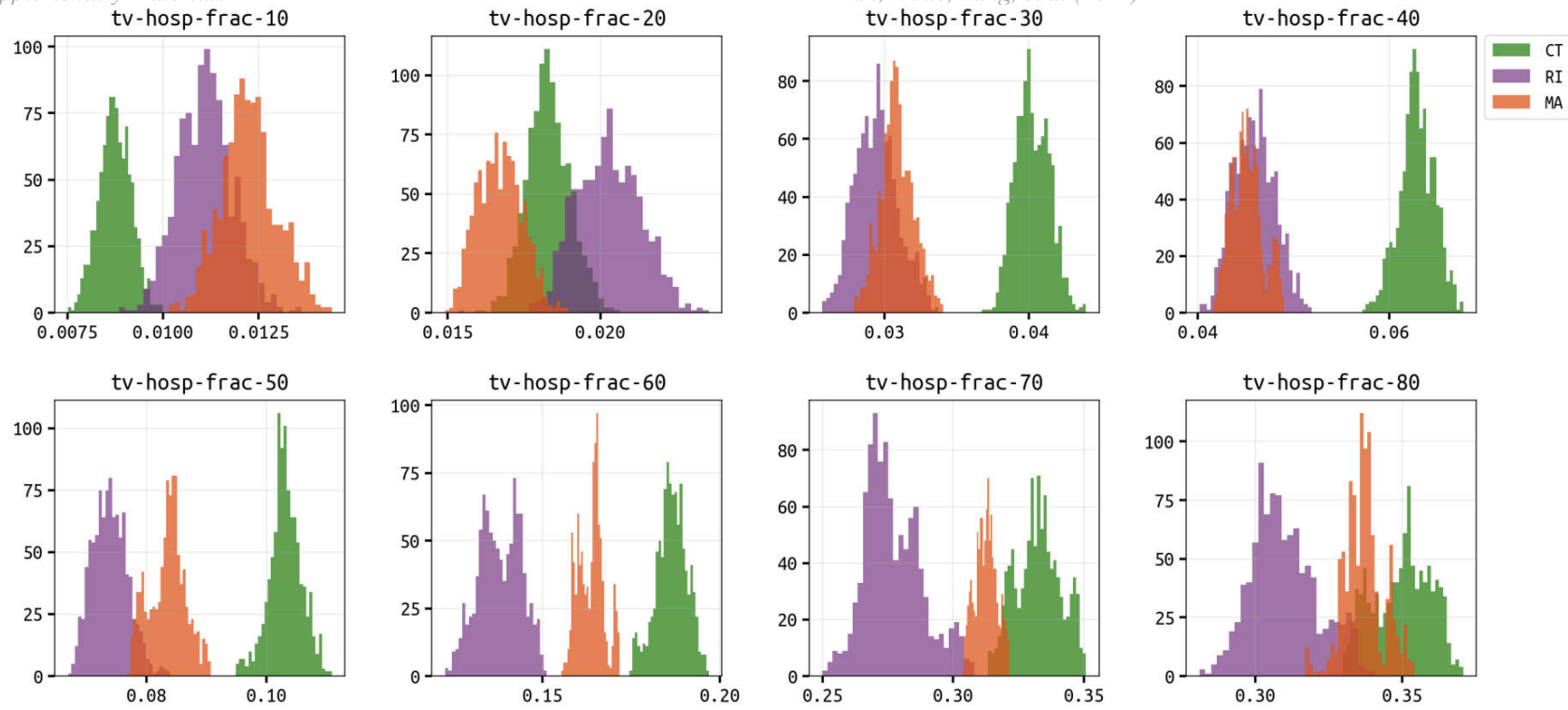

**Supplementary Figure 5.** Panels show posterior distributions for the probability of hospitalization for a symptomatic infection of SARS-CoV-2, by 10-year age band, starting at 10-19, 20-29, through to the 80+ age group. The 0-9 age group is assumed to have the same probability of hospitalization as the 10-19 age group.

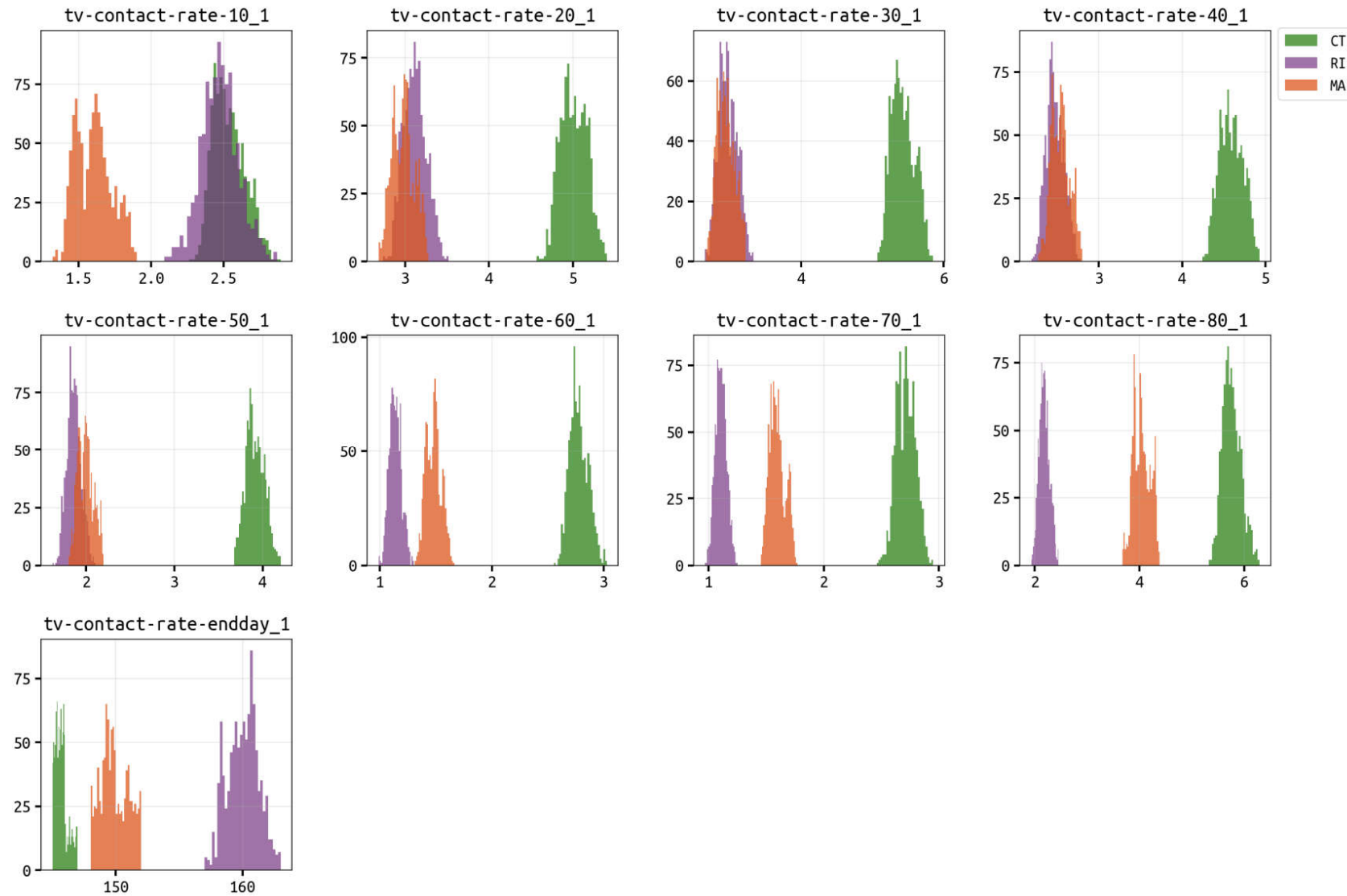

**Supplementary Figure 6.** Panels show posterior distributions for relative “transmission-capable contact rate” for the first period of inference which ends at **tv-contact-rate-endday\_1** (posteriors shown in bottom panel; day 150 is May 30 2020). Contact rates are broken down by age group (10-19, 20-29 to 80+) and are all presented as relative to the contact rate of the 0-9 age group.

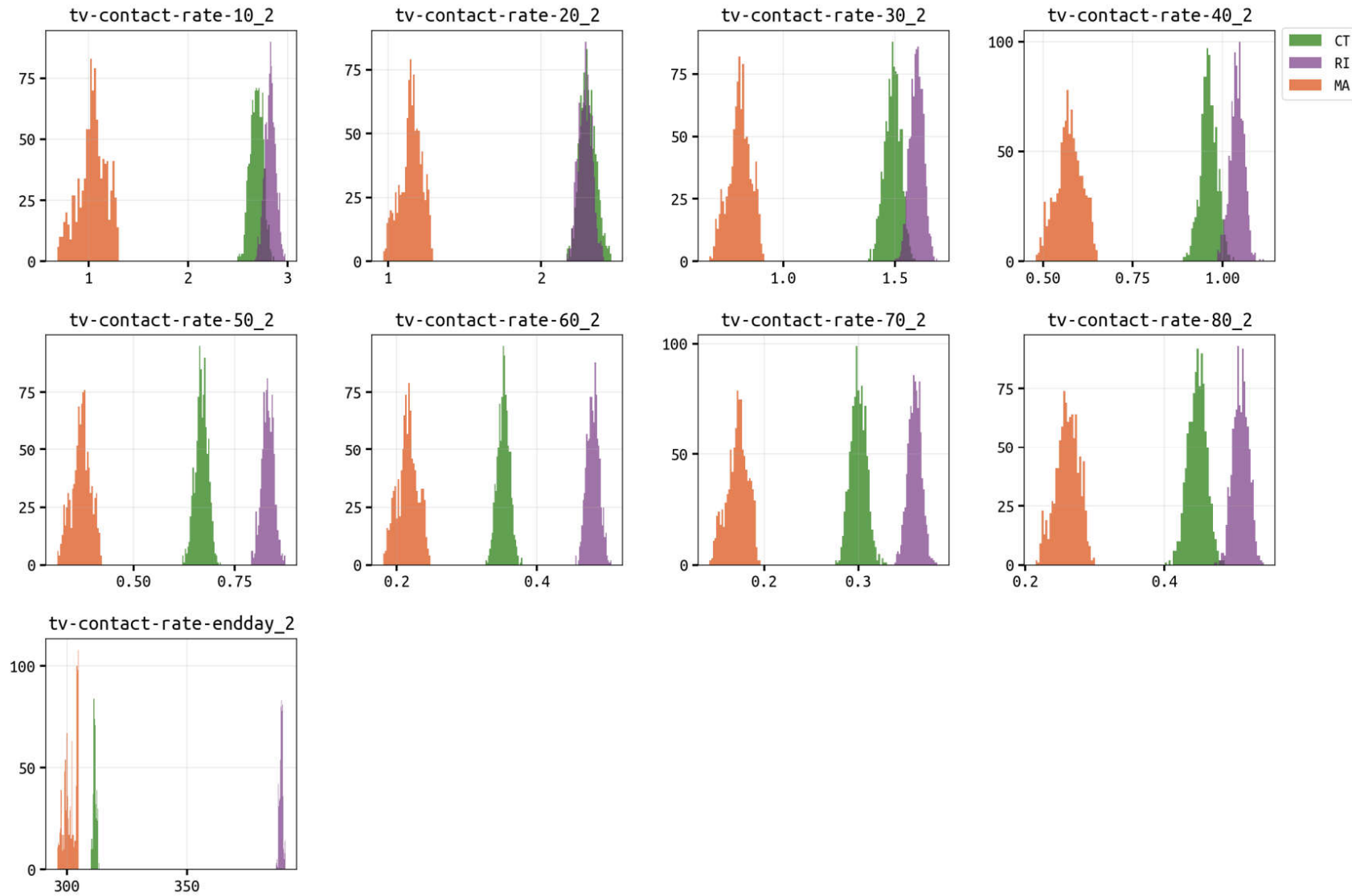

**Supplementary Figure 7.** Panels show posterior distributions for relative “transmission-capable contact rate” for the second period of inference which ends at **tv-contact-rate-endday\_2** (posteriors shown in bottom panel; day 300 is Oct 26 2020). Contact rates are broken down by age group (10-19, 20-29 to 80+) and are all presented as relative to the contact rate of the 0-9 age group.

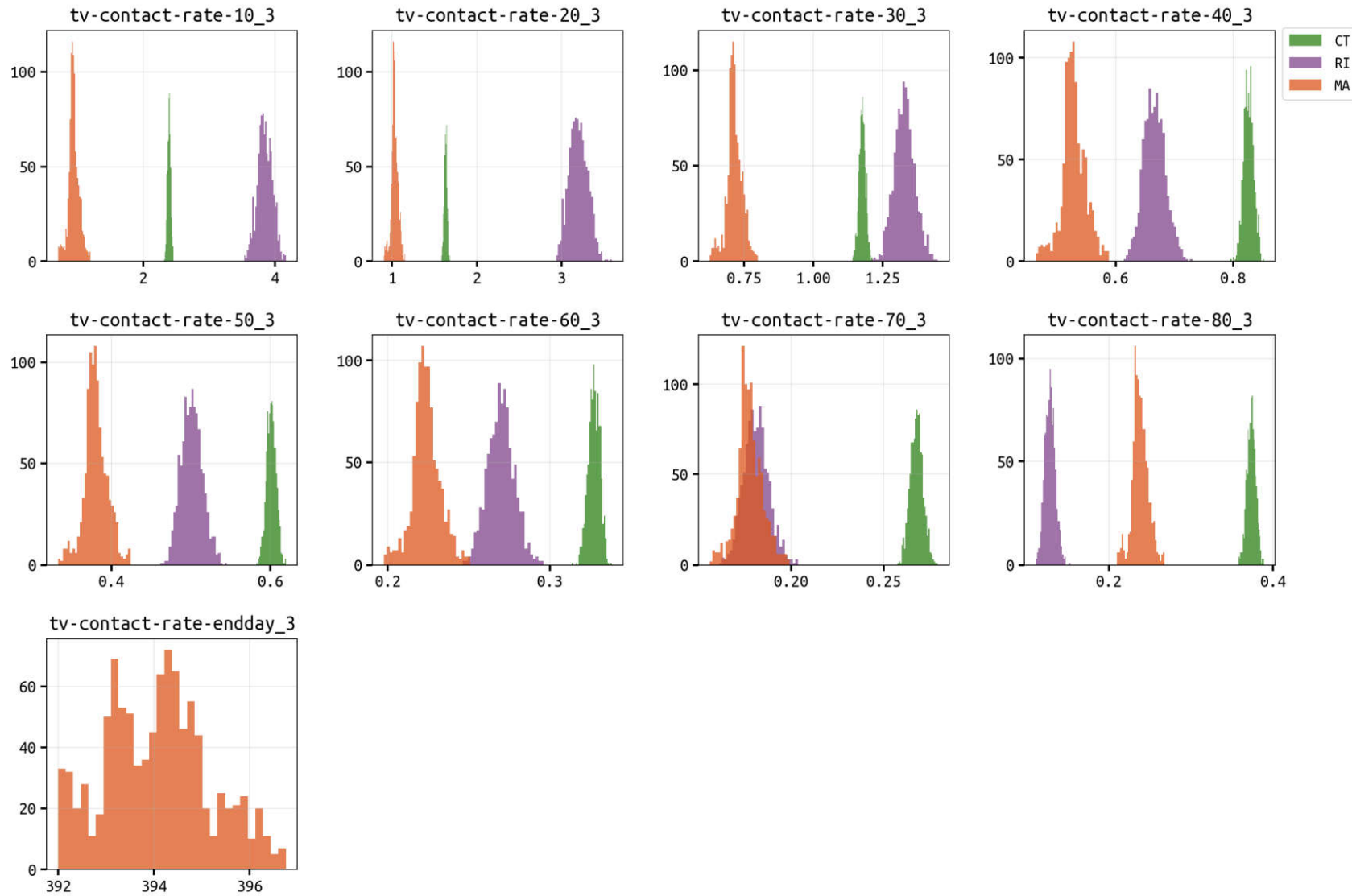

**Supplementary Figure 8.** Panels show posterior distributions for relative “transmission-capable contact rate” for the third period of inference which ends at **tv-contact-rate-endday\_3** for Massachusetts (posterior shown in bottom panel; day 394 is Jan 28 2021) or ends on May 31 2021 for Rhode Island and Connecticut. Contact rates are broken down by age group (10-19, 20-29 to 80+) and are all presented as relative to the contact rate of the 0-9 age group.

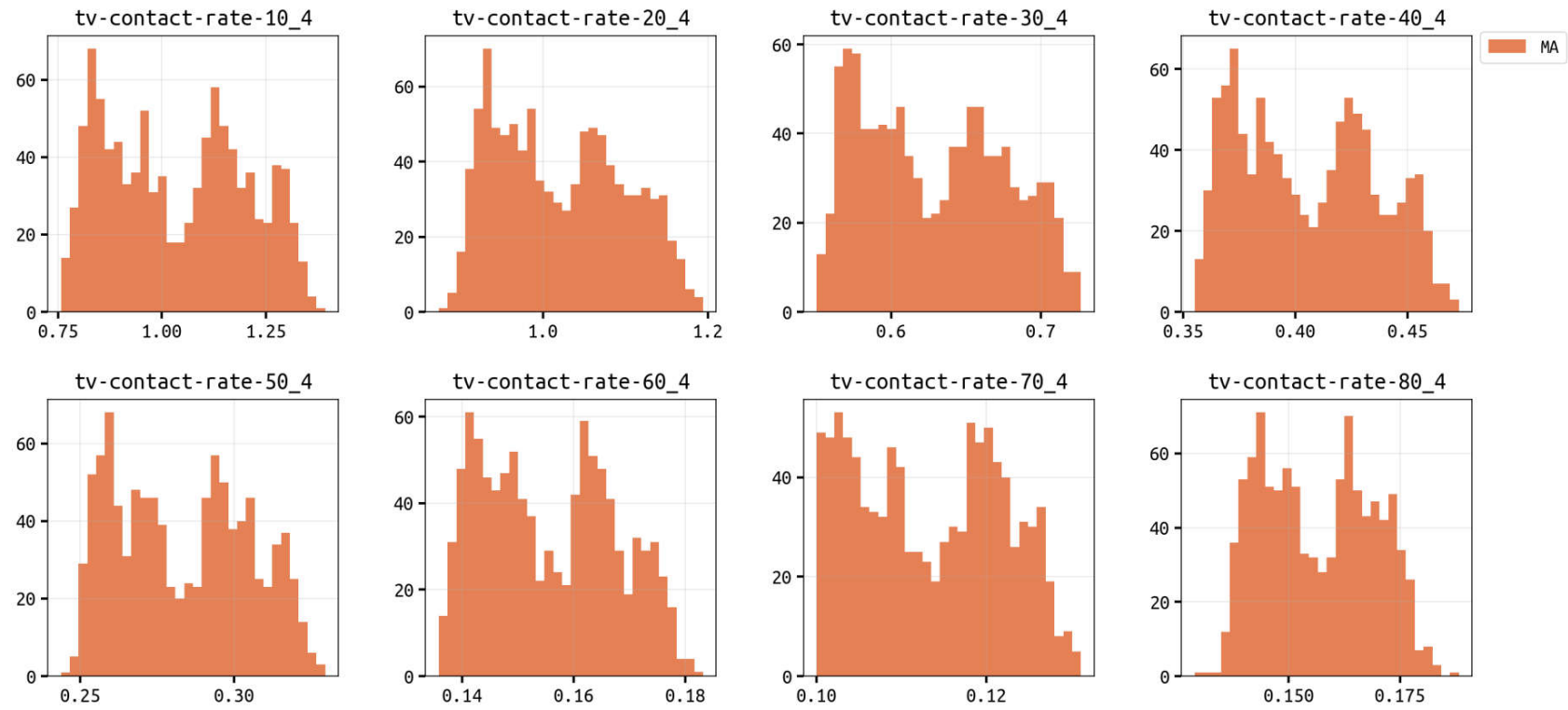

**Supplementary Figure 9.** Panels show posterior distributions for relative “transmission-capable contact rate” for the fourth period of inference which ends was only used for the Massachusetts inference (based on DIC and visual fit). Contact rates are broken down by age group (10-19, 20-29 to 80+) and are all presented as relative to the contact rate of the 0-9 age group.
